# Implications of *MGMT* promoter methylation and its downstream *hMSH2* mRNA in primary malignant glioma

**DOI:** 10.1101/19008763

**Authors:** Jeru-Manoj Manuel, K V L Narasinga Rao, G K Chetan

## Abstract

**Background:** Hypermethylation of 06-methylguanine DNA methyltransferase (*MGMT)*promoter seen in high grade gliomas (HGG) leads to the accumulation of O6-meG DNA damage which mispairs with thymine, requiring recognition by mismatch repair protein dimer MutSα, whose primary component is coded by Human MutS homolog protein 2 (*hMSH2)*. O6-meG repair necessitates the interaction/combined action of MGMT andhMSH2 to maintain genomic stability. Analysis of the correlation between *MGMT* methylation and *hMSH2*mRNAexpression in HGG and their role in the prognosis was explored.

**Methods:** Study was performed on 54 primary-frontal lobe HGG tumors, *MGMT* promoter methylation was detected by Q-MSP and Q-PCR was used to analyse *hMSH2* m-RNA expression levels.

**Results:** *MGMT* methylation was seen in 62%patients the mean percentage of methylation (PoM) being (17.62±17.20) %. *MGMT* PoM≥10% had improved Progression free survival (p=0.015) and ≥8% had better Overall survival (p=0.043), indicating its predictive significance. Over expression of *hMSH2* was seen in 50% patients with a median fold change of 2.74 (p=0.021). Univariate analysis of high *hMSH2* expression with therapy(CT+RT) showed poor PFS (p=0.002). There was no correlation between *MGMT* methylation and *hMSH2* expression.

**Conclusion:** *MGMT* PoM of ≥10% is a significant prognostic marker. Over expression of *hMSH2* is prognostic marker for poor treatment response. Lack of/aberrant correlation between *MGMT* and*hMSH2* could indicate impaired DNA repair of O6-meG in HGG, and this could be one of the factors responsible for both, gliomagenesis and variations in treatment response.

## 1. Background

Malignant high grade gliomas (HGG), are the fastest growing cancers, constitute Grade III and Grade IV tumors(Louis et al. 2007). Prognosis and survival for patients with HGG are poor, and tumors are highly resistant to chemotherapy and radiotherapy(Cabrini et al. 2015). Current standard of treatment includes chemotherapy (CT) with alkylating agents such as temozolomide (TMZ) along with intermittent radiotherapy(RT), but most patients’ exhibit rapid disease progression(Alvino et al. 2006).DNA damages per day occur at a high rate of 1000–1,000,000 molecular lesions per cell(H N et al. 2015).Hence, anefficient DNA repair system is required for the effective maintenance of genome integrity(Frosina 2009). Glioma develops through accumulation of genetic alterations due to DNA damage that allow the cells to escape normal growth-regulatory mechanisms(McLendon et al. 2008).DNA repair genes play a key dual role in primary gliomagenesis as well as treatment response (Lahtz and Pfeifer 2011; Perazzoli et al. 2015).DNA damaging agents often causes more than one type of damage, and more than one repair pathway may be involved in repair process of each DNA damage(Iyama and Wilson 2014; Kinsella 2009). Direct reversal of DNA damage and mismatch repair proteins are amongst the first to be recruited to the site of DNA damage, therefore expression of genes involved in it are key to maintain genomic stability(Iyama and Wilson 2014). Direct reversal of DNA damage repair system constitute,06-methylguanine DNA methyltransferase (*MGMT*) gene which codes for the suicide enzyme MGMT, that protects the cells against lesions caused by alkylation of DNA at the O-6 position of guanine.Methylation at CpG islands of the *MGMT* promoter leads to gene silencing and decreased expression, and is among the well-studied process in HGG (Manuel et al. 2016).*MGMT* promoter methylation status is a potential predictive marker for response to adjuvant radiotherapy plus chemotherapy, particularly with alkylating agents like temozolomide (TMZ)(Park et al. 2012; Villani et al. 2015).This epigenetic modification leads to cytotoxic effects, due to accumulation of errors in the form of O6-methylguanine (O6-meG)(Bearzatto et al. 2000).These damages induce the formation of O6-meG: T and O6-meG: C mispairs during DNA duplication and thereby subsequent recruitment of the DNA mismatch repair (MMR) system(Quiros, Roos, and Kaina 2010).MMR is a post-replicative functional downstream pathway to *MGMT*. It plays a ubiquitous role in ensuring replication fidelity and is responsible for correcting mismatch errors, preserving microsatellites’ and thereby genomic integrity(Modrich 1994). MMR can detect and repair base-base and insertion–deletion mismatches that are formed during DNA replication (Peltomaki 2003). Altered expression of MMR genes confer a mutator phenotype with small genetic disruptions, leading to emergence of microsatellite instability and somatic mutations(Vageli et al. 2012).MMR deficient tumor cells have accumulation of mutations coupled with replication errors. In accordance with ‘futile repair’ model, the MutS component of the MMR system recognizes and attempts to repair O6-meG:T andO6-meG:Cmispairs(Modrich and Lahue 1996). However, the repair event is not successful resulting in the degradation of the pyrimidine-containing strand followed by subsequent reinsertion of C or T opposite the O6-meG, since the methylated base is in the template strand and MMR targets the newly synthesized strand(Hsieh and Yamane 2008). Human MutS homolog protein 2 (hMSH2)is the primary component for mismatch recognition; it forms dimers MutSα along with hMSH6 or MutSβ along with hMSH3depending on the damage occurred (Fishel 2015). Hence, the optimum expression of *hMSH2* is critical among the other two genes for the entire MMR system to carry its function. Studies on exploring the functional implication of mismatch repair genes in HGG are limited, however there are reports indicating presence of microsatellite instability in adult high-grade gliomas, which is a sign of aberrant MMR functioning(Demokan 2006; Viana-Pereira et al. 2011).Persistent O6-meG mismatch errors are also associated with increased levels of sister chromatid exchanges and chromosome aberrations (Hsieh and Yamane 2008). These damages along with aberrant methylation are dependent on MMR and homologous recombination (HR) processing of the lesion(Kenyon et al. 2012).Recent evidence also suggests there are clinically relevant alterations in the DNA-MMR system beyond alterations in the *MGMT* gene (Stark et al. 2015). High MGMT enzyme activity is known to confer resistance to O6-G-methylating agents through detoxifying the DNA, but the actual apoptotic process requires a functional MMR system(Quiros, Roos, and Kaina 2010).Therefore, in the absence of MGMT activity due to methylation, cell sensitivity to O6-meG-methylating agents is largely dependent on MMR efficiency(Cabrini et al. 2015). Recognition of mismatch is the first step determining this efficiency(McFaline-Figueroa et al. 2015) and requires proper functioning of *hMSH2* mRNA. Given the crucial functional interactions, regulated levels of *MGMT* and MMR seem important for processing methylation directed DNA damages to maintain genomic stability (Fig 1).Therefore, exploring the correlation between *MGMT* and *hMSH2* would help in better understanding of HGG primarily due to the critical role of these two genes in gliomagenesis as well as for treatment response. In this context, the association between these genes and their impact on progression free survival (PFS) and overall survival (OS) in HGG were analyzed in this study.

**Fig1.**
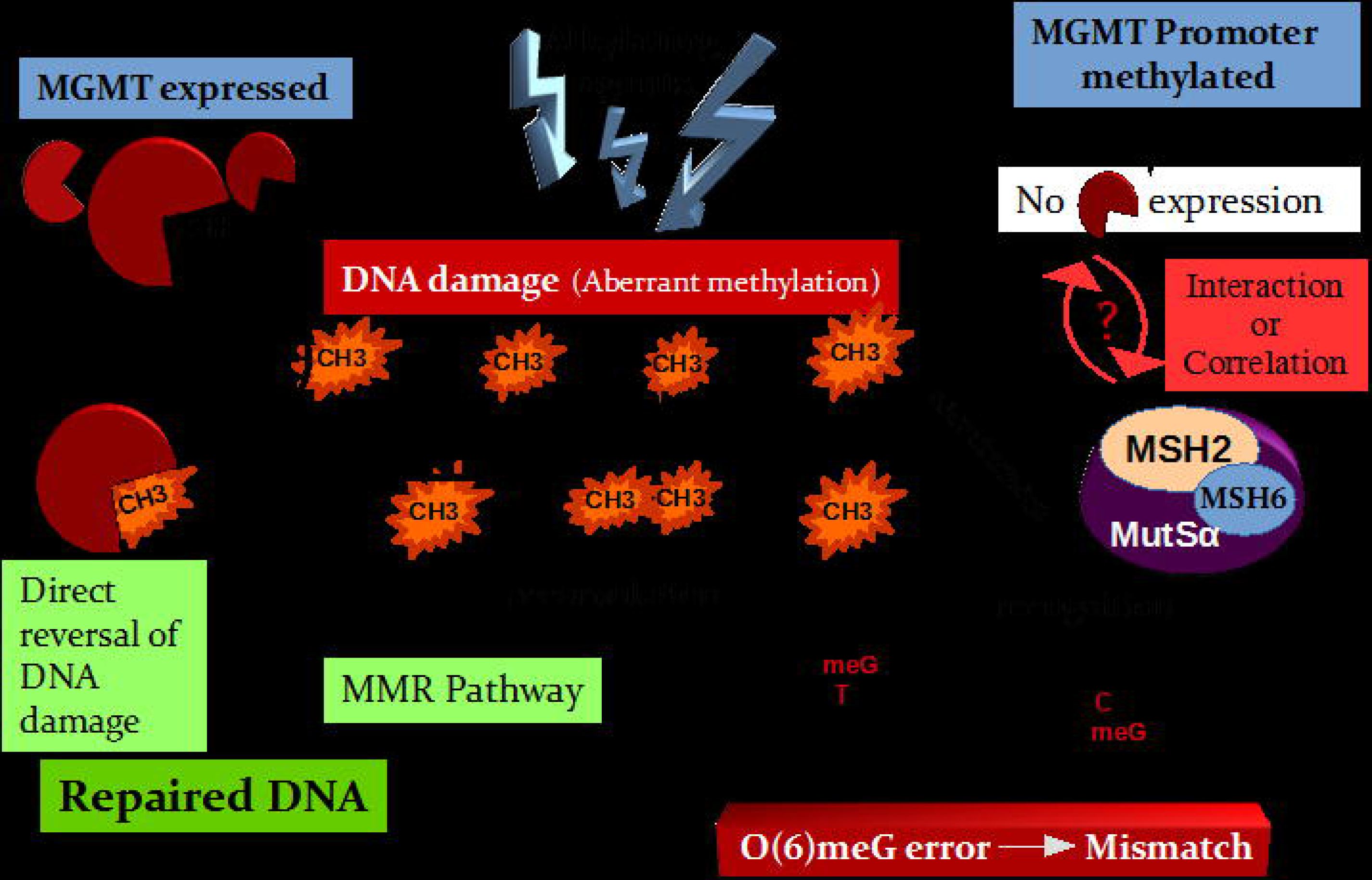
Graphical representation of the study hypothesis

## 2. Methods

### 2.1. Human patients

The resected frontal lobe primary HGG tumors were obtained from 54 adult (age 18-55 years) patients who underwent surgery at the Department of Neurosurgery, National Institute of Mental Health and Neurosciences (NIMHANS), Bangalore, India from December 2011-January 2014. Peripheral blood was also collected from the patients prior to surgery. Normal non-tumor cortical brain tissues (n=8) were collected from other patients who underwent surgery for non-glioma condition (frontal-lobe epilepsy). Prior informed consent was obtained from all the subjects as per the guidelines and approval of the Institute ethical committee. The resected tumor tissues were bisected, one portion was placed in RNAlater (Sigma Aldrich, Missouri, USA) and standard storage protocol was followed, till the isolation of nucleic acids. Remaining tissue was fixed in 10 % buffered neutral formalin, processed for paraffin section, and was used for histopathological grading, which was carried out at Department of Neuropathology, NIMHANS, Bangalore. Data on various demographic details such as age, sex, histological classification and post-surgery adjuvant treatment status were collected from medical records section. Pre and post-operative scans (MRI/CT) were analyzed at Department of Neuro-imaging and Radiology, NIMHANS, Bangalore to check for extent of resection. Patient follow-up evaluation was monitored for up to 24-months from date of surgery to check PFS and OS of the patients.

### 2.2. Isolation of Nucleic acids

Histologically characterized tissues which contained >95 % tumor cells were used for the analysis. DNA isolation from tissues and blood were carried out using Nucleospin® Tissue and Nucleospin® Blood L (Macherey Nagel GmbH & Co. KG, Germany), respectively.RNA isolation from both tumor and control brain tissues were carried out using Pure Link^™^ RNA Mini kit (Ambion, California, USA). DNA and RNA were quantified using Nanodrop ND 2000c (Thermo Scientific, Wilmington, USA). DNA and RNA samples having purity of 1.8–1.99 (A_260/280_) and >2.00 (A_260/280_), respectively were used.

### 2.3. Bisulphite conversion and real-time methylation specific PCR (Q-MSP)

Bisulphite conversion of DNA (500ng-1µg) was carried out using MethylCode^™^Bisulphite Conversion kit (Invitrogen, Carlsbad, CA, USA). Real time Methylation specific PCR (Q-MSP) was carried out using the eluted bisulphite modified DNA as template in 1:3 fold dilutions (1-2µL). Comparative Ct method was used and the PCR conditions were: 10 minutes at 95°C, then 45 cycles of 95°C for 15 seconds, 55°C for 15 seconds, and 60°C for 45 seconds in 10µL reaction volume set using 1× TaqMan Universal PCR Master Mix II without UNG (Applied Biosystems, Foster City, CA, USA). Methylation and unmethylation-specific primers and probes along with an endogenous control of *COL2A* (Rivera et al. 2009), were used. Singleplex experiment was performed in triplicates in a MicroAmp optical 96-well reaction plate (Applied Biosystems, Foster City, CA, USA). Each plate included non-template control (water), an unmethylated control (bisulfite treated normal leukocyte DNA), and a methylated control (bisulfite treated DNA from methylated glioma cell line U251-MG) along with the samples. The cell line was cultured in the central laboratory and tested in prior for its methylation status. To validate the Comparative Ct method, a relative standard curve experiment using a serial dilution of DNA from methylated glioma cell line U251, was used as the calibrator to check the amplification efficiency of both the target and housekeeping genes (Efficiency between 90-110% was considered optimal). Samples with a Ct-value above 35 were censored (resulting in a quantity of 0). The percentage of methylated reference (PMR) was calculated for each sample from the median quantity value from the triplicates by dividing the *MGMT*/*COL2A* quantity ratio in the target by the *MGMT*/*COL2A* quantity ratio in the methylated control (Rivera et al. 2009). Threshold value for scoring methylation positive samples was defined based on the qMSP result of the internal control bisulfite treated normal leukocyte DNA, all of which had PMR values of zero. Samples with a PMR value above zero were scored as methylation positive. The samples which had amplification in both methylated and unmethylated targets where considered as methylated based on the PMR value (≥1). The percentage of methylation (PoM) was calculated using (RQ=2^-ΔΔ*C*t^) *100, where ΔΔ*C*t =Δ*C*t *MGMT*-Δ*C*t Cal-U251.

### 2.4. Reverse transcription and Q-PCR

Total RNA (500ng) extracted from tissue samples (∼30mg) was reverse transcribed using 2X-with Oligo (dT) SuPrimeScript RT Premix (GENETBIO Inc., Daejeon, Korea). The optimized conditions in a final volume of 20μl were 5mins at 65°C, 60mins at 50°C followed by heating at 70°C for 10mins in an Eppendorf Personal Mastercycler (Hamburg, Germany) and the product was stored at -20°C.

Real-time PCR was carried out for *hMSH2* gene using cDNA template in 1:20 fold dilution. All quantitative PCR reactions were carried out in three replicates of each sample using a reaction volume of 10μl reaction containing 0.5μl of 20 x Assays-on-Demand^™^ Gene Expression Assay Mix (Applied Biosystems, Foster City, CA, USA; Assay ID-Hs00954125-m1; FAM™ dye labeled MGB probe). 2x TaqMan Universal PCR Master Mix II, no AmpErase/UNG (Applied Biosystems) were used and reactions were carried out in Applied Biosystems 7500 Fast Real-Time PCR System (Foster City, CA, USA). *18S rRNA* (Assay ID-Hs99999901_s1; FAM™ dye labeled MGB probe) was used as endogenous controls and each plate included ‘No template controls’ in triplicates for both the *hMSH2* and *18srRNA*. The thermal cycler was programmed for an initial denaturation step of 10min at 95°C followed by 45 thermal cycles of 15 sec at 95°C, 60 sec at 60°C. Fluorescent data were converted into cycle threshold (*C*t) measurements, and we calculated gene expression values by comparative threshold cycle (ΔΔ*C*T) method, which uses the formula 2^-^ΔΔ*C*t to calculate the expression of target genes normalized to a calibrator. *C*t data for *hMSH2*and *18srRNA*in each sample were used to create Δ*C*T values [Δ*C*T _Sample_ =*C*T_*hMSH2*_−*C*T_*18srRNA*_].Thereafter, ΔΔCt values were calculated by [Δ*C*T _Sample_ –Δ*C*T _Calibrator_], the Δ*C*T values of combined normal brain tissues were designated as calibrators. Relative quantity (RQ) of primary HGG compared with the non-glioma brain tissues was calculated with the equation: RQ= 2^−^ΔΔ*C*t. A relative standard curve experiment was performed to check the efficiency (90-110% was considered optimal) of the target and the housekeeping gene (Perez-Cabornero et al. 2009).

### 2.5. Statistical evaluation

Statistical analysis was carried out using IBM SPSS 20.0 software for Windows. Normality of distribution of data was checked using Shapiro-Wilk test. Non-parametric tests were used for those that were found to be not normally distributed. Optimal cut-off value for classifying *MGMT* PoM as high or low was estimated by using the Receiver Operating Characteristics (ROC) curve analysis (Villani et al. 2015).Comparisons between percentage of *MGMT* methylation and mRNA expression levels among the different grades and types were performed using the Kruskal-Wallis test. Kaplan Meier survival curves were used to check PFS and OS, univariate comparisons were made using log-rank test. Multivariate analysis to check independent prognostic value of variables was carried out using Cox proportional hazard mode. Probable correlation between *MGMT* promoter methylation and mRNA expression of *hMSH2* was analyzed using Spearman correlation test.

## 3. Results

### 3.1. Cohort characteristics and clinical features

Study group consisted of fifty four adult patients (Mean age: 39.2±9.9) with histologically confirmed frontal/fronto-insular HGG [Grade III:Anaplastic Oligodendroglioma (AOD), Anaplastic mixed Oligoastrocytoma (AOA), Anaplastic Astrocytoma (AA); Grade IV:Glioblastoma (GB)].For clinical end point analysis, extent of resection-gross total resection(GTR)or sub-total resection (STR) and adjuvant treatment following surgical procedure comprising of either radiotherapy (RT) plus chemotherapy with TMZ and/or no therapy was taken into consideration (Frequencies are as mentioned in Table 1). In this study group, 18 patients refused to go ahead with post-surgery therapy due to various socio-economic causes. [AOD-Anaplastic oligodendroglioma; AOA-Mixed anaplastic oligoastrocytoma; AA-Anaplastic Astrocytoma; GB-Glioblastoma; PFS-Progression free survival; OS-Overall survival; CT-Chemotherapy; RT-Radiotherapy]

**Table 1:** *MGMT* methylation status detected using PMR value (>1)

| Variables |  |  |  | Number of patients | MGMT Methylated samples[Frequency] |
| --- | --- | --- | --- | --- | --- |
| Total Number |  |  |  | 54 | 35[64.8%] |
| Age (years) |  | ≤39 |  | 29 | 22[75.8%] |
|  |  | >39 |  | 25 | 13 [44.8%] |
| Gender |  | Male |  | 40 | 25[62.5%] |
|  |  | Female |  | 10 | 10 [71.4%] |
| Grade and Type | III | Total |  | 40 | 29 [72.5%] |
|  |  | AOD |  | 27 | 23 [85.2%] |
|  |  | AOA |  | 7 | 4 [57.1%] |
|  |  | AA |  | 6 | 2 [33.3%] |
|  | IV | GB |  | 14 | 6 [42.9%] |
| PFS | Progressed<br>n=39 |  | G-III | 25 | 14[56%] |
|  |  |  | G-IV | 14 | 6[42.9%] |
|  | Not progressed<br>n=15 |  | G-III | 15 | 15[100%] |
|  |  |  | G-IV | 0 | -- |
| OS | Survived<br>n=31 |  | G-III | 30 | 29[96.7%] |
|  |  |  | G-IV | 1 | 1[100%] |
|  | Not Survived<br>n=23 |  | G-III | 10 | 5[50%] |
|  |  |  | G-IV | 13 | 6[46.2%] |
| Post-surgery adjuvant therapy | CT+RT<br>n=38 |  | G-III | 33 | 24[72.7%] |
|  |  |  | G-IV | 5 | 3[60%] |
|  | No<br>n=16 |  | G-III | 7 | 5[71.4%] |
|  |  |  | G-IV | 9 | 3[33.3%] |

[AOD- Anaplastic oligodendroglioma; AOA- Mixed anaplastic oligoastrocytoma; AA-Anaplastic Astrocytoma; GB-Glioblastoma; PFS-Progression free survival; OS-Overall survival; CT-Chemotherapy; RT-Radiotherapy]

### 3.2. Q-MSP

Q-MSP analysis based on PMR valuation revealed positive *MGMT* promoter methylation in 62% (35/54) of the samples. Positive methylation statuses among the different variables are given in Table1, with an overall median PoM of 10.16% (ranging from 0.9-64). The mean *MGMT* PoM for the methylated sampleswas17.62±17.20% and this was used for further analysis (Fig 2). Statistically significant differences were observed in PoM for grade III (p=0.05) and oligodendroglioma (p=0.015) (Table.2).

**Table 2:**
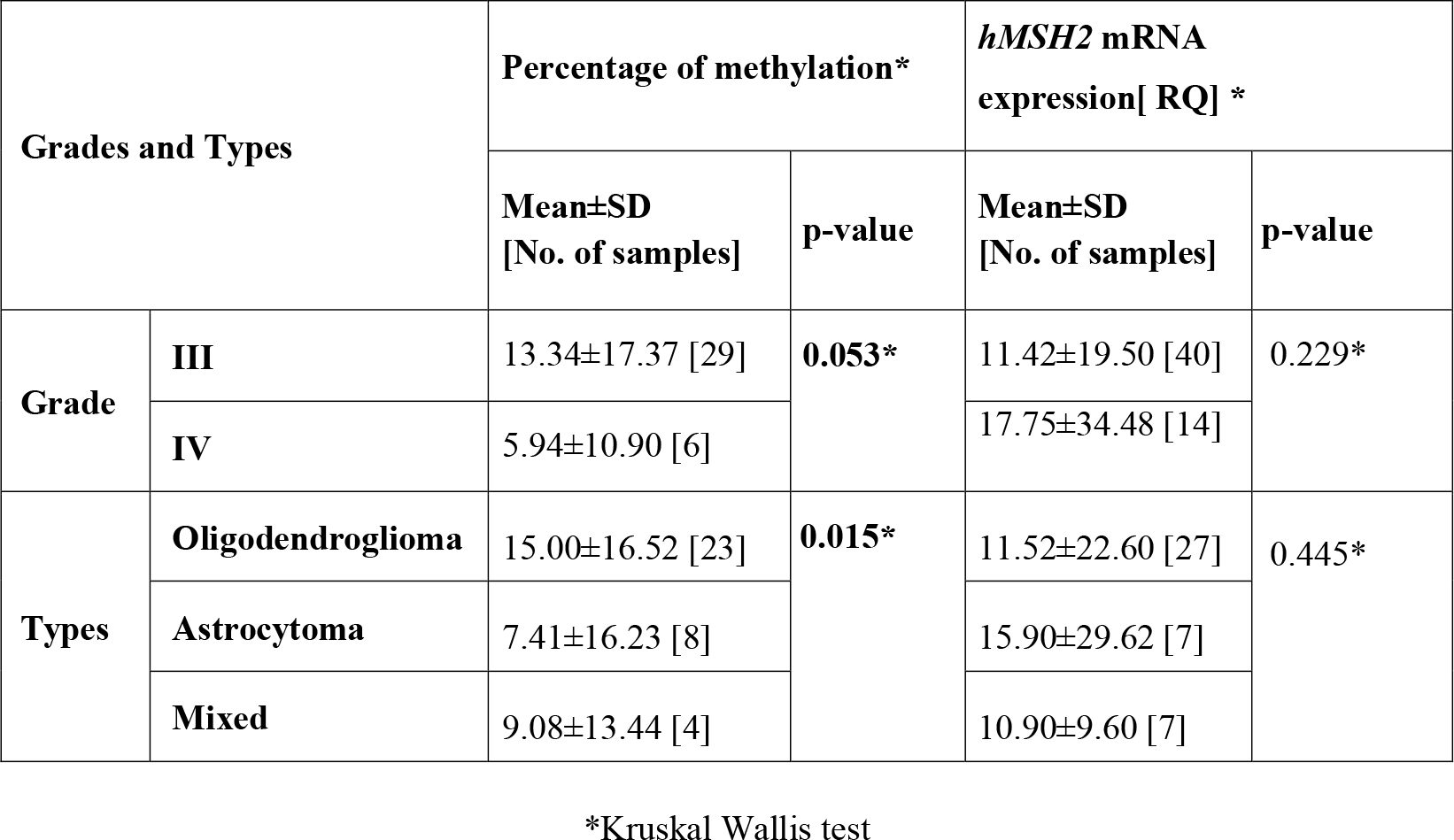
*MGMT* Percentage of methylation [PoM] and *hMSH2* mRNA expression across grade and type

| Grades and Types |  | Percentage of methylation* |  | *hMSH2* mRNA expression[ RQ] * |  |
| --- | --- | --- | --- | --- | --- |
| | | Mean±SD<br>[No. of samples] | p-value | Mean±SD<br>[No. of samples] | p-value |
| Grade | III | 13.34±17.37 [29] | 0.053* | 11.42±19.50 [40] | 0.229* |
| | IV | 5.94±10.90 [6] | | 17.75±34.48 [14] | |
| Types | Oligodendroglioma | 15.00±16.52 [23] | 0.015* | 11.52±22.60 [27] | 0.445* |
| | Astrocytoma | 7.41±16.23 [8] | | 15.90±29.62 [7] | |
| | Mixed | 9.08±13.44 [4] | | 10.90±9.60 [7] | |

\*Kruskal Wallis test

**Fig2.**
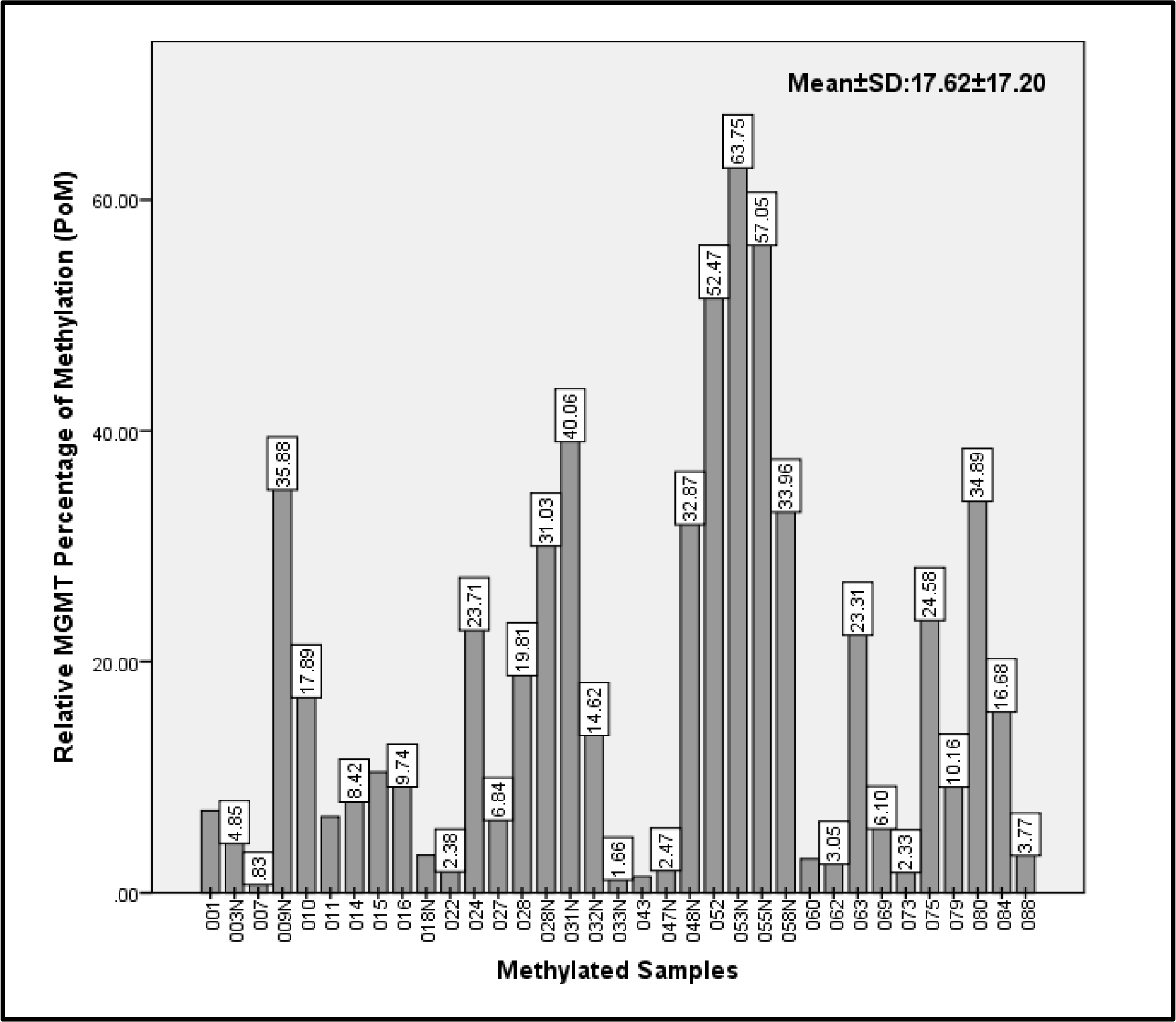
Bar graph depicting *MGMT* PoM across the 35 methylated samples

### 3.3. Receiver Operating Characteristics Analyses

The best criteria for *MGMT* PoM that predicted PFS and OS were 10% and 6%, respectively, the findings of Area under curve (AUC) are presented in Table 3. When cut-off value of 10% MGMT methylation was chosen, a sensitivity of 72% and specificity of 73% in predicting PFS (*p*=0.006) was observed. Whereas, a cut-off value of 8% of MGMT methylation ensured a sensitivity of 65% and specificity of 67% in predicting OS (*p*=0.043).

**Table 3:** Receiver operating characteristics (ROC) analysis of *MGMT* PoM-cutoff value for predicting PFS and OS

|  | **PFS** | **OS** |
| --- | --- | --- |
| **AUC\* [95% CI]** | 0.744 (0.613-0.875) | 0.662 (0.514-0.810) |
| **PoM cut-off** | ≥10% | ≥8% |
| **Sensitivity** | 72% | 65% |
| **Specificity** | 73% | 67% |
| **p-value** | **0.006** | **0.043** |

\*AUC- Area under curve

### 3.4. Expression of *hMSH2*mRNA

Expression of *hMSH2*mRNAin HGG relative to non-glioma brain tissue revealed over-expression (median was considered as cut-off)in 50% of subjects (Fig 3a),with median fold change of 2.74 (p=0.021).Overall mean *hMSH2* mRNA expression level observed was 13.06±24.07.There were differences in the levels of*hMSH2*expressionbetween the grades and types, but not statistically significant; Grade IV and astrocytoma were seen to have higher expression (Table 2).

**Fig3a.**
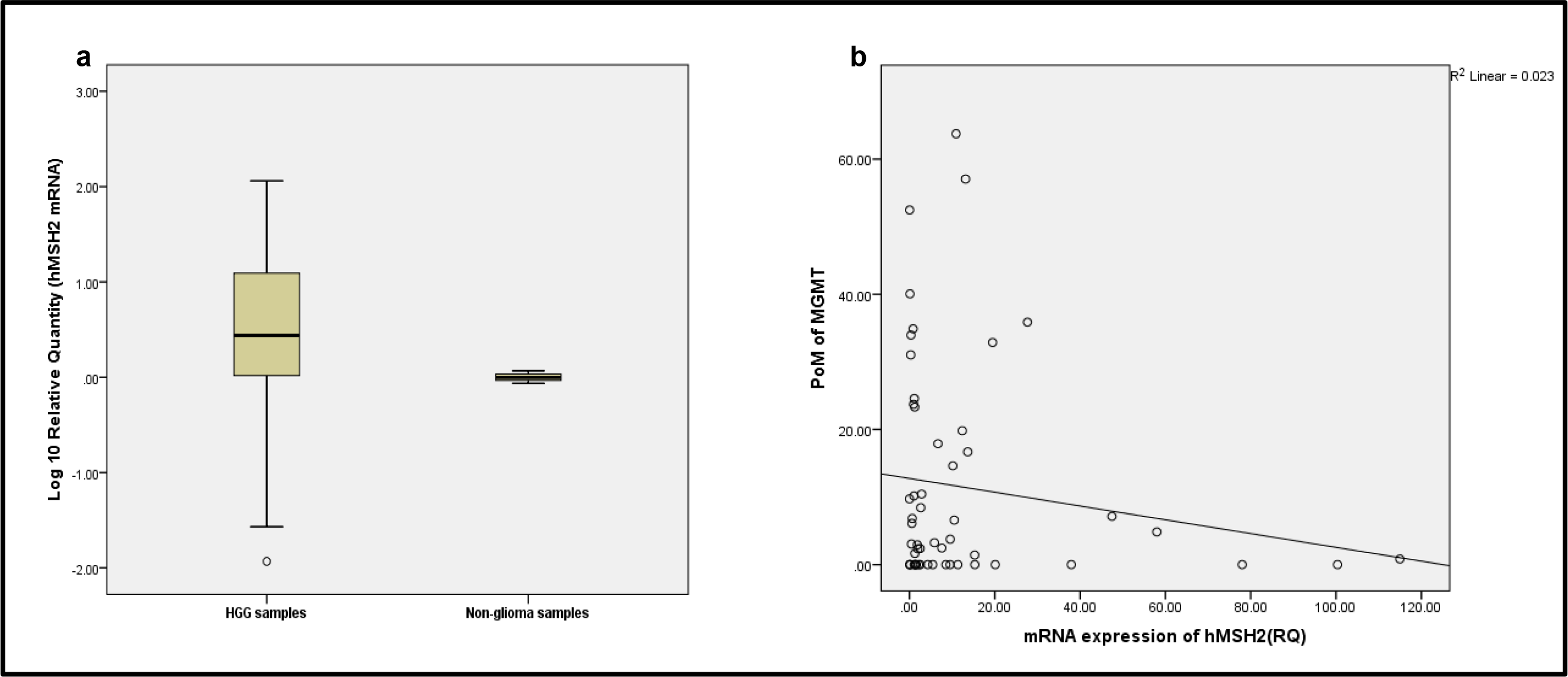
Box plot depicting *hMSH2* mRNA expression, HGG samples are over expressed relative to non-glioma tissue (p=0.021; Median-2.74fold difference) **3b**Scatter plot depicting aberrant inverse correlation between *MGMT* POM and *hMSH2* mRNA expression (ρ=-0.183)

### 3.5. Correlation between *MGMT* and *hMSH2*

Association between the statuses’ of *MGMT* methylation and *hMSH2* expression using chi-square analysis was checked and no significant association[χ^2^=0.081; (p=0.776)] was observed. Further, Spearman’s correlation analysis between *MGMT* PoM and *hMSH2* mRNA expression (Fig 3b) also showed a negative correlation [ρ=-0.183;p=0.294] which was statistically not significant.

### 3.6. Survival analysis

Univariate analysis using Kaplan-Meier survival and log-rank tests showed that *MGMT* methylation, age ≤39, grade III, oligodendroglioma and gross total resection (GTR)were good survival indicators for both PFS and OS (Table 4). PFS and OS was checked separately for both the grades.

**Table 4:** Univariate survival analysis demonstrating impact of basic patient characteristics on PFS and OS

| Variables |  | PFS |  | OS |  |
| --- | --- | --- | --- | --- | --- |
| | | Mean ± SD (months) | p-value | Mean ± SD ( months) | p-value |
| $\leq 39$ years | | 18.24±1.3 | **0.001** | 21.68±1.3 | **<0.0001** |
| >39 years | | 10.10±1.6 | | 14.42±1.7 | |
| III | AOD | 18.90±1.3 | **<0.0001** | 21.70±1.0 | **<0.0001** |
| | AOA | 16.15±2.2 | | 22.58±1.6 | |
| | AA | 16.50±2.8 | | 21.96±1.6 | |
| IV | GBM | 4.22±0.8 | | 7.58±1.4 | |
| GTR (n=27) | | 20.40±1.0 | **<0.0001** | 23.34±0.5 | **<0.0001** |
| STR (n=27) | | 8.52±1.3 | | 13.30±1.70 | |

AOD-Anaplastic Oligodendroglioma, AOA- Anaplastic Oligoastrocytoma, AA- Anaplastic Astrocytoma, GBM- Glioblastoma; GTR-Gross total resection, STR- Sub-total resection

#### 3.6.1. PFS

In the 24months of follow-up 72.2% patients had HGG progression. *MGMT* PoM greater than 10% showed good PFS (p=0.015). Mean PFS for samples with ≥10%*MGMT* methylation was 18.15±1.88 months and for <10% *MGMT* methylation it was 12.30±1.38 months (Fig 4a). *MGMT* methylation status had a significant impact on PFS (p=0.001) in grade III, but not in grade IV tumors (p=0.527). Univariate *hMSH2*gene expression did not show any statistically significant association with PFS or OS. However, univariate analysis combining the two variables*hMSH2* expression status along with adjuvant therapy showed significant impact on PFS in grade III tumors(p=0.002). Patients who underwent therapy (CT+RT) showed a difference in the median PFS, which was 17 months for patients with high *hMSH2*and24 months for those with low *hMSH2* expression (Figure 5 a & b). Interestingly, patients who did not undergo therapy had almost similar median of 11 and 12 months for high and low expression respectively. Grade IV tumors also demonstrated a similar trend as grade III, but was not statistically significant (p=0.099) (Table 5).

**Table 5:** Impact of therapy, *MGMT* methylation and*hMSH2* expression on PFS and OS

| Variables |  | PFS |  | OS |  |
| --- | --- | --- | --- | --- | --- |
|  |  | Mean ± SD<br>(months) | p-value | Mean ± SD<br>(months) | p-value |
| Adjuvant Therapy |  |  |  |  |  |
| GIII | CT+RT | 19.60±1.1 | <0.001 | 23.10±0.5 | 0.001 |
|  | No | 11.88±1.9 |  | 15.18±2.9 |  |
| GIV | CT+RT | 6.60±1.2 | 0.049 | 11.20±2.2 | 0.032 |
|  | No | 2.88± 0.9 |  | 5.56±1.4 |  |
| MGMT Methylation status |  |  |  |  |  |
| GIII | M | 20.55 ±1.1 | 0.001 | 22.14±1.0 | 0.040 |
|  | UM | 11.46±1.3 |  | 21.06±1.5 |  |
| GIV | M | 5.17±1.1 | 0.527 | 8.83±1.7 | 0.952 |
|  | UM | 3.50±1.2 |  | 6.62±2.0 |  |
| hMSH2 expression |  |  |  |  |  |
| GIII | High | 17.40±1.5 | 0.730 | 21.60±1.2 | 0.552 |
|  | Low | 18.70±1.5 |  | 22.14±1.1 |  |
| GIV | High | 3.85±1.2 | 0.950 | 7.28±2.1 | 0.677 |
|  | Low | 4.58±1.3 |  | 7.85±1.8 |  |
| GIII | High & therapy* | 18.12±1.6 | 0.002 | 22.77±0.9 | 0.007 |
|  | High& No therapy | 13.33±2.8 |  | 15.00±4.0 |  |
|  | Low &therapy | 21.26±1.2 |  | 23.44±0.5 |  |
|  | Low& No therapy | 11.00±2.6 |  | 14.25±4.6 |  |
| GIV | High & therapy | 8.00±2.0 | 0.099 | 11.33±3.4 | 0.098 |
|  | High & No therapy | 2.20±0.4 |  | 4.25±1.8 |  |
|  | Low & therapy | 7.00±1.0 |  | 11.00±3.0 |  |
|  | Low& No therapy | 3.60±1.5 |  | 6.6±2.2 |  |

\*Therapy- CT+ RT; M-Methylated; UM-Unmethylated

**Fig4.**
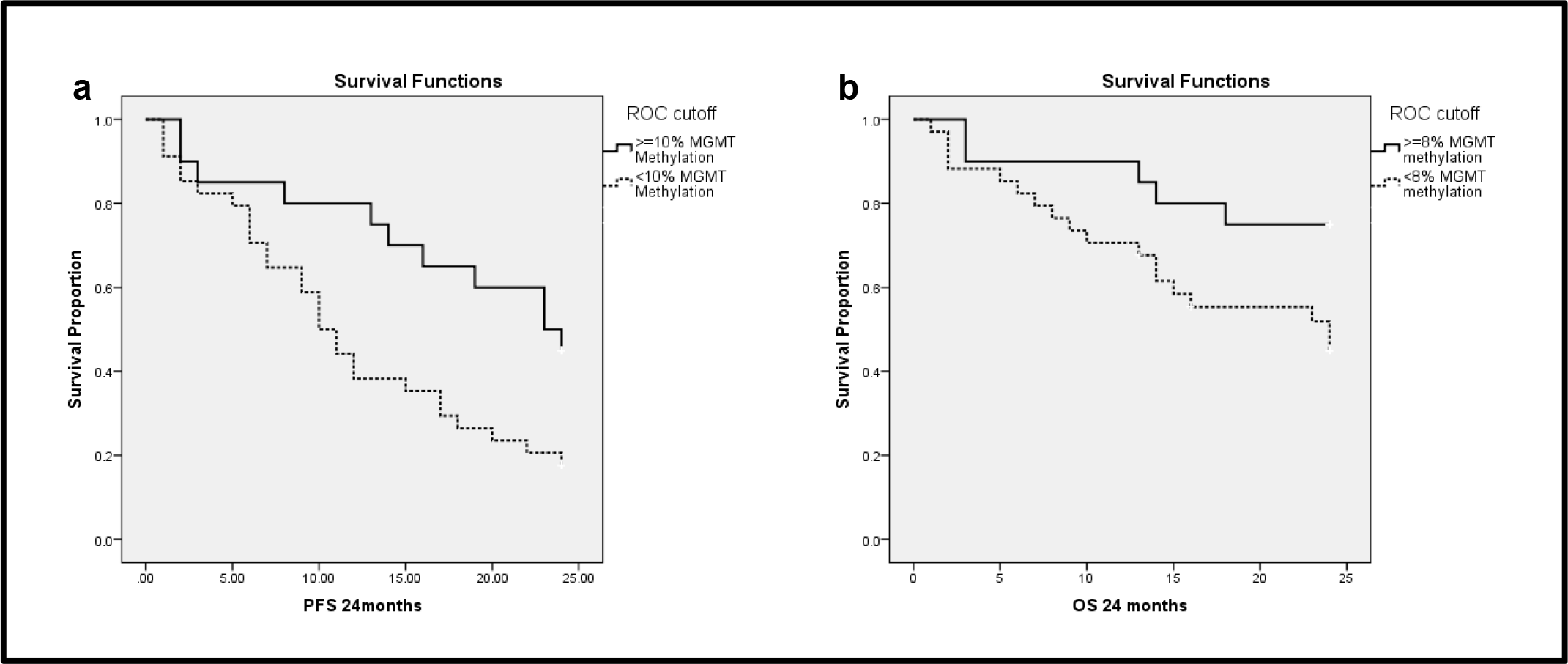
Kaplan Meier Survival analysis demonstrating the efficacy of **4a**PoM of 10% MGMT methylation in predicting PFS (p=0.015) **4b**PoM of 8% MGMT methylation in predicting OS (p=0.043)

**Fig5.**
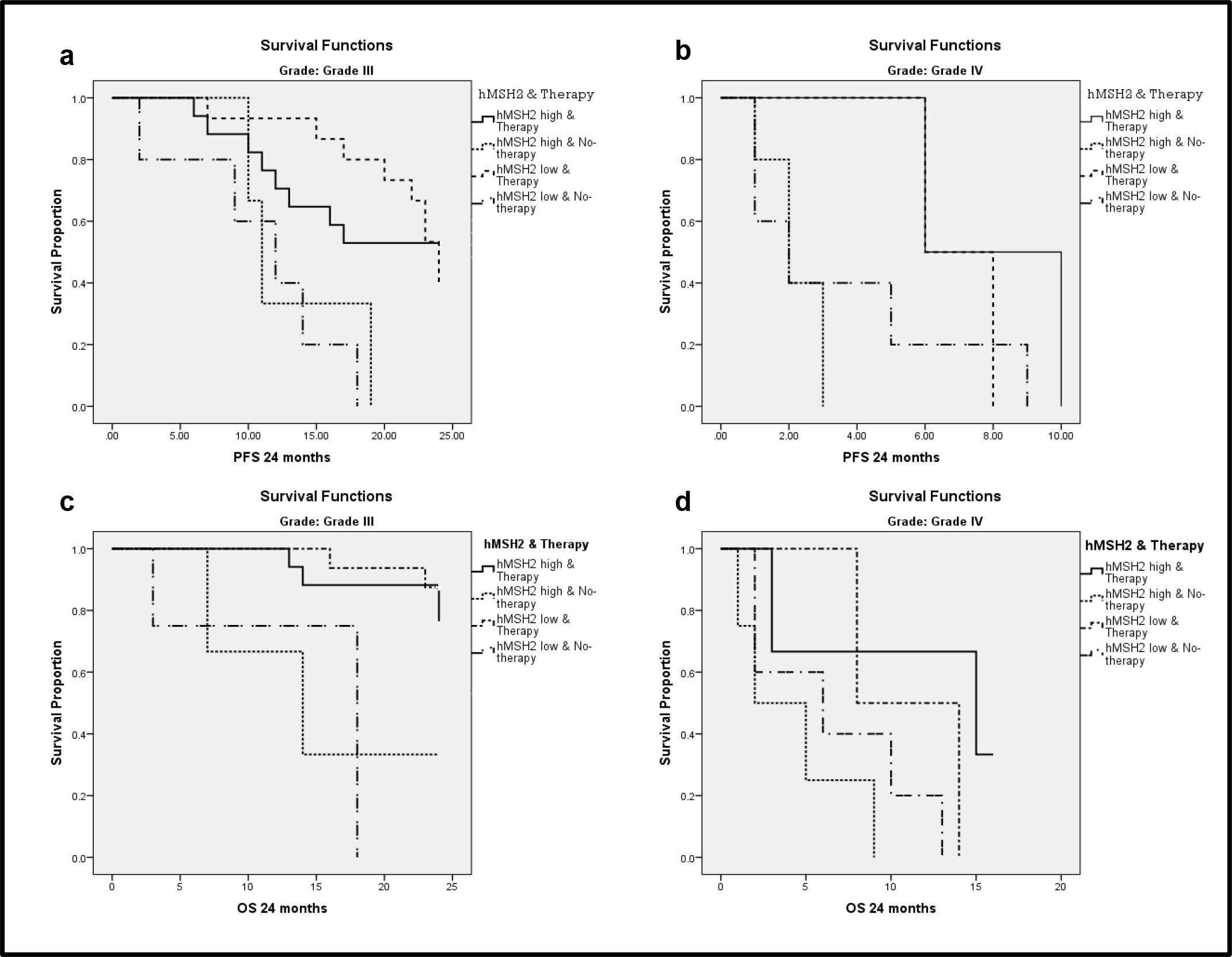
Univariate survival analysis showing combination of *hMSH2* expression and therapy status (CT+RT), its impact on survival; *hMSH2* low expression when adjuvant therapy was/was not administered has better survival when compared to *hMSH2* high expression when adjuvant therapy was/was not administered **5a** Grade III tumors impact on PFS (p=0.002) **5b** Grade IV tumors impact on PFS (p=0.099) **5c**Grade III tumors impact on OS (p=0.007) and 5**d** Grade IV tumors impact on OS (p=0.098)

#### 3.6.2. OS

In the 24 months of follow-up 42.6% patients did not survive. *MGMT* PoM greater than 8% showed good OS (p=0.043). Mean OS for ≥8%*MGMT* methylated samples was 20.55±1.50 months and for <8% *MGMT* methylated samples was 16.98±1.49 months (Fig 4b). *MGMT* methylation status did not seem to affect OS in grade IV tumors (p=0.952), but had a significant impact in grade III tumors (p=0.040). Similar to PFS, a significant impact of *hMSH2*expression combined with adjuvant therapy was seen on OS (Fig 5 c & d) in grade III (p=0.007) and a trend towards significance was observed in grade IV (p=0.098).

Multivariate analysis using Cox proportional hazard revealed age ≤39, grade III, GTR, adjuvant therapy and *MGMT* methylation were good independent prognostic indicators for PFS. The same variables with the exception of MGMT methylation were found to be significant for OS as well (Table 6).

**Table 6:**
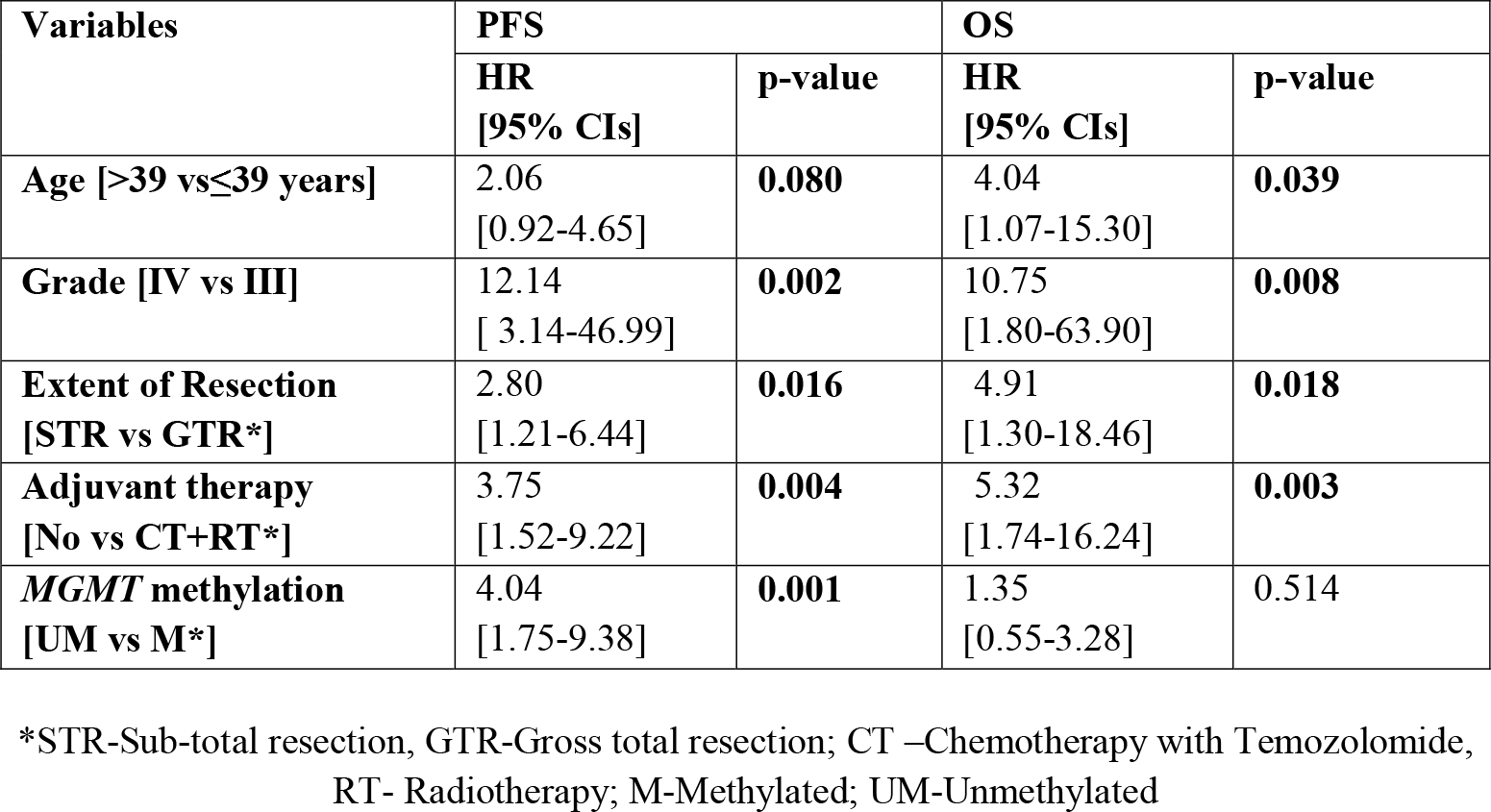
Multivariate Cox proportional hazards models predicting PFS and OS

## 4. Discussion

Differences in the ability to repair DNA damage contributes to the cancer risk as well as treatment response (Demokan 2006). Methylation of *MGMT* at 0-6-position remains unrepaired as there are no other known repair enzymes which carry out the function(Liu et al. 2009); leading to accumulation of single base pair mismatches(Pukkila et al. 1983).This DNA damage response pathway is of importance due to its rolein gliomagenesis and also for determining efficacy of therapy using alkylating agents(Rodríguez-hernández et al. 2013).Processing of O6-meG is dependent on methylation status of *MGMT* promoter region (Warren, Forsberg, and Beese 2006) and on MMR proteins(Zhang et al. 2010).Given the exploratory nature of our investigation and this being the first study to check the relationship between *MGMT* promoter methylation and gene expression of its functional downstream gene *hMSH2* in HGG, we did not have a specific directional hypothesis.

*MGMT* methylation is known to be a good prognostic and predictive indicator which is supported by reports(Manuel et al. 2016; Sciuscio et al. 2011). However, information about the extent of *MGMT* methylation is required for better understanding of its role in influencing expression of other genes involved in functional downstream pathways(Iliadis et al. 2012). qMSP was used in this study to determine PoM due to the specificity of the method and also helps in analyzing more number of CpG sites(Parrella et al. 2009).PoM was found to be significantly higher in grade III when compared with grade IV tumors, suggesting that the extent of *MGMT* methylation is dependent on grade and could be another factor for progression of severity in glioma. De-methylation of *MGMT* leading to decrease in PoM may be a factor responsible for progression of tumor grade, along with many other molecular events. A similar mechanism may apply to the type of glial cell as well. we observed oligodendroglioma to have significantly higher methylation and better prognosis when compared with astrocytoma, which is in accordance with literature wherein oligodendroglioma is seen to have higher methylation and better prognosis (Mo et al. 2005).This could probably indicate that higher *MGMT* methylation may be one of the factors responsible for providing oligodendroglioma the prognostic advantage. PoM of ≥10% *MGMT* methylation was seen to be a significant prognostic indicator for PFS and ≥8% for OS, this observation is consistent with earlier reports (Håvik et al. 2012; Villani et al. 2015). We also observed the independent efficacy of *MGMT* methylation in predicting PFS through multivariate analysis, as reported in other studies (Qiu et al. 2014; Rivera et al. 2009).

*MGMT* methylation leads to accumulation of mismatch errors(Qiu et al. 2014), that has to be corrected by the MMR system. If there are MMR aberrations especially in the mismatch recognition dimer MutSα,it would lead to accumulation of mutations, thereby increasing replication errors and genomic instability (Modrich and Lahue 1996).hMSH2is critical among the three MutSαproteins for the efficient functioning of mismatch recognition (McFaline-Figueroa et al. 2015).José L. McFaline-Figueroa et al. 2015, demonstrated that a minor change in hMSH2 protein expression has major impact on response of glioblastoma to TMZ(McFaline-Figueroa et al. 2015). However, studies on *hMSH2* mRNA expression are sparse, and there is no data available on the levels of mRNA expression in primary HGG in the literature. It is important to understand the expression of *hMSH2* owing to its’ evolutionary significance and ubiquitous function in post DNA replication and repair. Differential DNA MMR expression indicates mutation rate variations(Supek and Lehner 2015),and*hMSH2*is a critical gene for efficient MMR. However, our results interestingly demonstrate an equal distribution of over and low*hMSH2*mRNA expression, suggesting the underlying aberrant expression status in HGG, in accordance with a report on hMSH2 protein levels being aberrant (Alkam et al. 2013).Differences in expression levels between the grades and cell types correlating with severity were also observed (Table 3). Grade II and III glioma is known to have decreased *hMSH2* expression when compared with grade IV (Zheng et al. 2006). Similarly, in our study higher *hMSH2*expression was seen in grade IV and as astrocytoma, which probably indicates the increased underlying mutations, a characteristic in these sub-types of HGG.

Based on the functions of these genes, a positive correlation between *MGMT* methylation and *hMSH2* expression is essential for efficient repair of O^6^-meGs and for maintaining genomic stability. Higher *MGMT* methylation would lead to *MGMT* gene silencing and accumulation of O6-meG(Park et al. 2012),this could induce an increase in*hMSH2* expression for carrying out repair caused due to deficiency of MGMT enzyme(Frosina 2009). On the other hand, in HGG we found a negative correlation between extent of *MGMT* methylation and levels of *hMSH2*mRNA expression i.e., higher the methylation of *MGMT*, lower the expression of *hMSH2*.This probably indicates the lack of regulation between these two genes in HGG tissues and implicates a dysfunctional repair of O^6^-meG.There are no reports in the past indicating this kind of association between genes of two repair mechanisms; however, some studies have examined the correlation of methylation status with mRNA and protein expression within a single repair mechanism. *MGMT* and *hMSH2* methylation is seen to have positive direct correlation with its own mRNA and protein expressions, respectively (Alkam et al. 2013; Uno et al. 2011). Conversely, this impaired correlation between *MGMT* and *hMSH2*could be one of the early events which may be responsible for increasing the mutations across the HGG genome. Since, studies have shown that defects in the mismatch repair system result in accelerated accumulation of mutations in critical genes and lead to, progression to malignancy (Demokan 2006).However, it is difficult to ascertain the speculated aberrant correlation due to the complexity of biological relationship between promoter methylation and its impact on gene regulation (Sciuscio et al. 2011), especially in a primary HGG tissue. HGG is known to have resistance to CT and RT, especially therapy with alkylating agents(Weller et al. 2009).*MGMT* and MMR are major determinants of the tumor response to TMZ with concomitant RT (Zhang et al. 2010). Knowledge of *MGMT* levels and MMR status in cancer cells might be useful predictors of their chemotherapeutic response (Casorelli, Russo, and Bignami 2008). This is demonstrated in our study, wherein *MGMT* methylation and *hMSH2* low expression along with CT+RT was seen to have a better prognosis. There were patients in our cohort who did not receive any form of adjuvant therapy and therefore difference in survival time between them and patients who underwent CT+RT were considered for analysis. The observed statistically significant prognostic impact enabled us to determine the affectivity of CT+ RT, in terms of extending the survival for few months. Over-expression of *hMSH2*mRNA was seen to have poor PFS when CT+RT are administered. This could be due to *hMSH2* interfering with the TMZ sensitivity, wherein the DNA damage caused by TMZ may be repaired by an increased*hMSH2*expression.

## 5. Conclusion

DNA damages are known to occur ubiquitously, the DNA repair mechanisms individually or in combination target and repair the damages. This coordinated function is highly essential in determining the genomic stability and thereby in mediating normal cellular physiology. Failure DNA repair systems are known to be associated with all cancers, but it’s not well explored in glioma. Studies so far have explored mutations and protein expression of individual DNA repair mechanisms. However, a better approach would be to analyze the influence of mutations/errors in one DNA repair mechanism on another, as their interdependence or lack of it could underlie complex conditions like glioma. Future studies taking this aspect into consideration are necessary, evidenced by our study which provides preliminary indications of the importance of analyzing correlation between two DNA repair mechanisms with a similar biological function. The observations also emphasises the importance of analysing *hMSH2* along with *MGMT* methylation status and warrants further research.

## Data Availability

The first author of this manuscript is responsible for the possession of data referred to in the manuscript.

## Acknowledgement

Ph.D program of the author Jeru-Manoj Manuel and part of the study were funded by the University Grants Commission (UGC-MANF: No.F1-17.1/2011/MANF-CHR-KAR-2143/ SA-III/Website), New Delhi. This study was also funded partly by the grants from Department of Science and Technology-Science and Engineering Research Board (DST-SERB), New Delhi (No. SR/SO/HS-233/2012).Authors thank patients and their relatives who consented to participate in the study. Neuropathologists at NIMHANS are acknowledged for histopathological classification of the glioma tissues. Authors wish to specially thank Dr. Vijay K Kalia (Professor, Dept. of Biophysics NIMHANS) and his students Ms. Kalyani Thakur and Mr. Sai Shyam V (Dept. of Biophysics, NIMHANS) for providing the cultured methylated U251-MG cell line. The services of Dr. Venkatesh H N and Dr. Debarati Ghosh in helping out with sample collection and technical assistance, and Dr. Sibin M K for analytical support, are also acknowledged.

## Declaration

## Abbreviations

HGG: (High grade glioma)
O-6meG: (DNA adduct at O-6 position)
MGMT: (O-6-Methylguanine DNA methyltransferase)
MMR: (Mismatch repair genes)
hMSH2: (human MUTS homolog 2)
AOD: (Anaplastic oligodendroglioma)
AOA: (Anaplastic mixed Oligoastrocytoma)
AA: (Anaplastic astrocytoma)
AE: (Anaplastic ependymomas)
GB: (Glioblastoma)
PMR: (Percentage methylated reference)
PoM: (Percentage of Methylation)
PFS: (Progression free survival)
OS: (Overall survival)
CT: (Chemotherapy)
RT: (Radiotherapy)
TMZ: (Temozolomide)

## Authors Contributions

JMM carried out the sample collection, genetic studies, conception and design, acquisition of data, interpretation of results, drafting and revising the manuscript. CGK and NRKVL conceived of the study, and participated in its design coordination and helped to draft the manuscript, agree to be accountable for all aspects of the work in ensuring that questions related to the accuracy or integrity of any part of the work are appropriately investigated and resolved and have given final approval of the version to be published.

## Ethical approval

All procedures performed in studies involving human participants were in accordance with the ethical standards of the institutional and/or national research committee and with the 1964 Helsinki Declaration and its later amendments or comparable ethical standards. The sample collection was approved by the Institute ethical committee of NIMHANS (NIMH/DO/ETHICS SUB-COMMITTEE/2014).

## Informed consent to publish

After explaining the details of the study, prior informed consent was obtained from all individual participants; they were also informed that the results obtained will be used for publication.

